# Metformin use and the risk of incident breast cancer: a systematic review and meta-analysis accounting for time-related biases, and why imeglimin cannot yet be evaluated

**DOI:** 10.64898/2026.11.01.26367001

**Authors:** Misuzu Akieda, Yuta Horita, Hiroyoshi Harada, Masayoshi Mori, Yusuke Murata, Kenji Ohe

## Abstract

**Background:** Observational studies have suggested that metformin lowers breast cancer risk, but many were susceptible to immortal time and time-window bias. Imeglimin, a structurally related glucose-lowering drug approved in Japan in 2021, has been proposed as a candidate for cancer prevention.

**Methods:** We searched PubMed (1 November 2026) and the reference lists of earlier meta-analyses for cohort and case-control studies that reported breast cancer-specific estimates for metformin use and incident breast cancer. One report per source population was pooled with a DerSimonian–Laird random-effects model. Studies were classified by whether they addressed time-related biases, using a new-user design, time-varying exposure or target trial emulation. For imeglimin, we applied the same criteria in a systematic search of PubMed and ClinicalTrials.gov, assessed whether a meta-analysis was feasible, and reviewed non-clinical and structural data.

**Results:** The search identified 368 PubMed records and 28 reports from other sources. Twenty-two reports from 15 source populations were eligible, and 12 independent studies were pooled. The pooled relative risk (RR) was 0.89 (95% CI 0.74–1.07; I^2^ = 90.5%). In nine studies that addressed time-related biases, the RR was 1.00 (0.94–1.08; I^2^ = 0%). In three that did not, or were unclear, it was 0.69 (0.56–0.84); the between-group difference was Q = 12.78, p = 0.0004. The difference was not significant when the one unclear study was reclassified (Q = 1.15, p = 0.28). For imeglimin, the search returned 218 PubMed records and 10 registered studies, but no clinical study reported breast cancer or any cancer outcome. No meta-analysis was possible. The available evidence consisted of one in vitro study in breast cancer cells, one docking study, and models of other cancers at concentrations at least about 40-fold above clinical plasma levels. A comparative cohort would need about 631 breast cancer events to detect a hazard ratio of 0.80.

**Conclusions:** In studies that addressed time-related biases, metformin use was not associated with incident breast cancer. A meta-analysis of imeglimin and breast cancer is not yet possible, because no clinical study exists and exposure time since approval is short. Its structural relation to metformin does not by itself predict a preventive effect. The next step should be an adequately powered new-user, active-comparator cohort study.

**Registration:** PROSPERO CRD420261301586 (submitted 6 February 2026; registered 1 November 2026). Deviations from the protocol are reported in the Methods.

## Introduction

Breast cancer is the most frequently diagnosed cancer in women worldwide [1]. Type 2 diabetes is associated with a modestly increased risk of breast cancer, and hyperinsulinaemia and insulin-like growth factor signalling have been proposed as mediating pathways [2, 3]. Metformin lowers hepatic glucose output and circulating insulin, inhibits mitochondrial respiratory complex I, and activates AMP-activated protein kinase in experimental systems, which has made it a candidate for cancer chemoprevention [3].

The hypothesis that metformin reduces cancer incidence originated in an observational study of people with diabetes [4]. Meta-analyses of observational studies of breast cancer incidence have reached different conclusions: a 2012 meta-analysis reported a protective association limited to postmenopausal women with diabetes [5], whereas a 2018 GRADE-assessed meta-analysis found no significant association [6], and a 2023 meta-analysis again reported a lower risk [7]. A recurring explanation for such discrepancies is time-related bias, in particular immortal time bias and time-window bias, which systematically favour the exposed group when exposure is misclassified in time [8, 9]. In the adjuvant setting, the MA.32 randomised trial found no improvement in invasive disease-free survival with metformin in non-diabetic women with early breast cancer [10]; that trial addressed recurrence, not primary prevention.

Imeglimin, the first tetrahydrotriazine glucose-lowering agent, was approved in Japan in 2021 [11, 12]. It shares a guanidine-derived scaffold with metformin but has a distinct mitochondrial profile [11]. No clinical study has examined imeglimin and breast cancer incidence.

We therefore performed a systematic review and meta-analysis of primary studies of metformin use and incident breast cancer, with a comparison, planned before data extraction, of studies that did and did not address time-related biases. We then considered, from its chemical structure and known pharmacology, what can and cannot be inferred about imeglimin.

## Methods

### Protocol and reporting

This review is reported according to PRISMA 2020 [13]. A protocol was submitted to PROSPERO on 6 February 2026 and registered on 1 November 2026 (CRD420261301586). The registered protocol had a broader scope: metformin and imeglimin in cancer prevention and longevity. The present report deviates from it in six ways:

1. the outcome was restricted to incident breast cancer, and longevity was not examined;
2. animal studies were not included;
3. the search used PubMed, reference lists and targeted searches, not all the databases listed in the protocol;
4. no language restriction was applied;
5. study authors were not contacted for missing data;
6. certainty of evidence was not graded with GRADE.

The eligibility criteria for this report, the choice of effect measure and the subgroup analysis by handling of time-related bias were defined during the re-analysis, before data were extracted for this report, and are reported here in full.

### Eligibility criteria

- **Population:** adult women, with or without type 2 diabetes.
- **Exposure:** metformin use (ever, any or new use).
- **Comparator:** non-use of metformin among people with diabetes, initiation of another glucose-lowering drug (active comparator), or women without diabetes (sensitivity analysis only).
- **Outcome:** incident breast cancer. Studies of recurrence, survival, mortality, stage or tumour characteristics after diagnosis were excluded.
- **Designs:** cohort studies, nested case-control and case-control studies, and secondary analyses of randomised trials that reported a breast cancer-specific hazard ratio (HR), odds ratio (OR) or relative risk with a 95% confidence interval (CI). Reviews and meta-analyses were not pooled; their reference lists were used to identify primary studies.
- Studies reporting only all-cancer incidence, or only dose–response estimates without an ever-use or new-use contrast, were excluded.

### Information sources and search

PubMed was searched on 1 November 2026, without date or language limits, with the following strategy: (metformin[MeSH Terms] OR metformin[tiab] OR biguanide*[tiab]) AND (“breast neoplasms”[MeSH Terms] OR “breast cancer*”[tiab] OR “breast carcinoma*”[tiab] OR “breast neoplasm*”[tiab]) AND (incidence[tiab] OR risk[tiab] OR cohort*[tiab] OR “case-control”[tiab] OR “new user*”[tiab] OR “target trial”[tiab] OR epidemiolog*[tiab]). In addition, we screened the reference lists of earlier meta-analyses [5–7, 14, 15] and ran targeted searches for studies of metformin and cancer incidence that report breast cancer as one of several sites, because these often do not mention breast cancer in the title or indexing. Embase and Ichushi-Web were not searched. Titles and abstracts were screened by one reviewer, and every record retained at this stage was assessed against the eligibility criteria using the full text or, where the full text was unavailable, the abstract.

### Study selection and data extraction

For each study we extracted the first author, year, country, data source, design, comparator, effect measure, the most fully adjusted estimate with its 95% CI, and the method used to define exposure over time. Estimates were taken from the abstract or the full text, and the location of each number was recorded (Supplementary Table S2). When a study reported sulfonylurea versus metformin, the estimate was inverted (1/HR; CI limits inverted and swapped). Data were extracted by one reviewer assisted by a large language model (Claude, Anthropic). A second large language model (ChatGPT, OpenAI), which had no access to the first extraction, extracted the same items independently from the full-text PDFs using a standardised form and instructions. These instructions required the estimate to be copied exactly as printed, in the direction reported, with its location and a verbatim snippet. Every value used in the analyses was then checked against the original report, and all discrepancies were resolved by a human author (KO) against the source document. The extraction form, instructions and the record of discrepancies are provided in Supplementary Data.

### Handling of overlapping populations

Several reports used the same database. For the main analysis we kept one report per source population: for UK CPRD/GPRD, the target trial emulation by Dickerman et al. [16]; for Taiwan’s National Health Insurance data, the full-population cohort by Tseng [17]; and for the US Veterans Affairs system, the report with more breast cancer events (Abdallah et al. [18]). The other reports from these databases were used in sensitivity analyses.

### Classification of time-related bias

Each study was classified as having addressed time-related biases (Y) if exposure was defined at cohort entry in new users or initiators, if it was modelled as time-varying from a defined start of follow-up, or if a target trial was emulated. Studies were classified as not having addressed these biases (N) if earlier methodological work identified immortal time, time-window or time-lag bias in them [9], or if exposure was classified as ever versus never over the whole follow-up without a time-varying analysis. Studies in which the handling could not be determined from the available text were classed as unclear and grouped with N.

### Risk of bias

Risk of bias was assessed with the Newcastle–Ottawa Scale (NOS) for cohort and case-control studies [37], using operational rules fixed before scoring. For cohort studies, the selection items were awarded for a population-based or health-system cohort (S1), non-exposed women drawn from the same population (S2), exposure ascertained from prescription, dispensing or claims records (S3), and exclusion of prior cancer (S4). Comparability was awarded for adjustment for age (C1) and for body mass index or obesity (C2). The outcome items were awarded for ascertainment by cancer registry, record linkage or validated codes (O1), mean or median follow-up of at least 3 years (O2), and complete or quantified follow-up (O3). Equivalent rules were used for case-control studies. Totals of 7–9 were rated high, 4–6 moderate and 0–3 low quality. Each judgement was recorded with the supporting text from the report (Supplementary Table S3). Scoring was done by one reviewer assisted by a large language model and checked by a human author (KO). Because the NOS does not assess immortal time, time-window or time-lag bias, these were classified separately, as described above. A sensitivity analysis was restricted to high-quality studies.

### Synthesis

Hazard ratios, rate ratios and odds ratios were treated as approximations of the same relative risk (RR), because breast cancer is a rare outcome over the follow-up periods studied. Log RRs were pooled with a DerSimonian–Laird random-effects model [19]. As a sensitivity analysis we used restricted maximum likelihood with the Hartung–Knapp–Sidik–Jonkman correction. Heterogeneity was quantified with Cochran’s Q, I^2^ and τZ, and a 95% prediction interval was calculated. Differences between subgroups were tested with the Q test for between-group heterogeneity. Pre-planned subgroups were time-related bias handling (Y vs N), comparator (active vs non-use) and region. Sensitivity analyses included leave-one-out analysis, alternative choices among overlapping reports, inclusion of all reports, and inclusion of studies with non-diabetic comparators. Small-study effects were examined with a funnel plot and Egger’s regression test. Analyses were run in Python 3 (NumPy, SciPy), and the code and data are provided in Supplementary Data.

### Imeglimin: search and assessment of feasibility

We applied the same eligibility criteria to imeglimin: clinical studies of any design that reported incident breast cancer, or malignancy by site, in imeglimin users compared with a defined comparator. On 1 November 2026 we searched PubMed for “imeglimin” alone and combined with cancer terms (cancer OR neoplasm* OR tumor* OR tumour* OR malignan* OR carcinoma*) and with “breast”, and searched ClinicalTrials.gov for registered studies of imeglimin. Non-clinical studies of imeglimin in cancer cells or models were recorded and described, but were not eligible for pooling. We also estimated, using Schoenfeld’s formula [36], how many breast cancer events a comparative cohort study would need to detect a plausible effect.

## Results

### Study selection and characteristics

The PubMed search returned 368 records (Figure 1). After title and abstract screening, 27 reports were assessed in full. Of these, 13 were excluded and 14 were included. The exclusions comprised five reports with only a dose, duration or other non-ever-use contrast, three with no breast cancer-specific estimate, two with an ineligible design (case-only or cross-sectional), one on an outcome other than incidence, one duplicate cohort, and one case-control study whose estimates were internally inconsistent and could not be interpreted. A further 28 reports were identified from reference lists and targeted searches. Of these, eight were included, 15 were excluded, and the breast cancer estimate could not be obtained for five (Ruiter 2012, van Staa 2012, Ferrara 2011, the RECORD and ADOPT trials, and Ngwana); these are listed as awaiting assessment (Supplementary Table S4).

**Figure 1.**
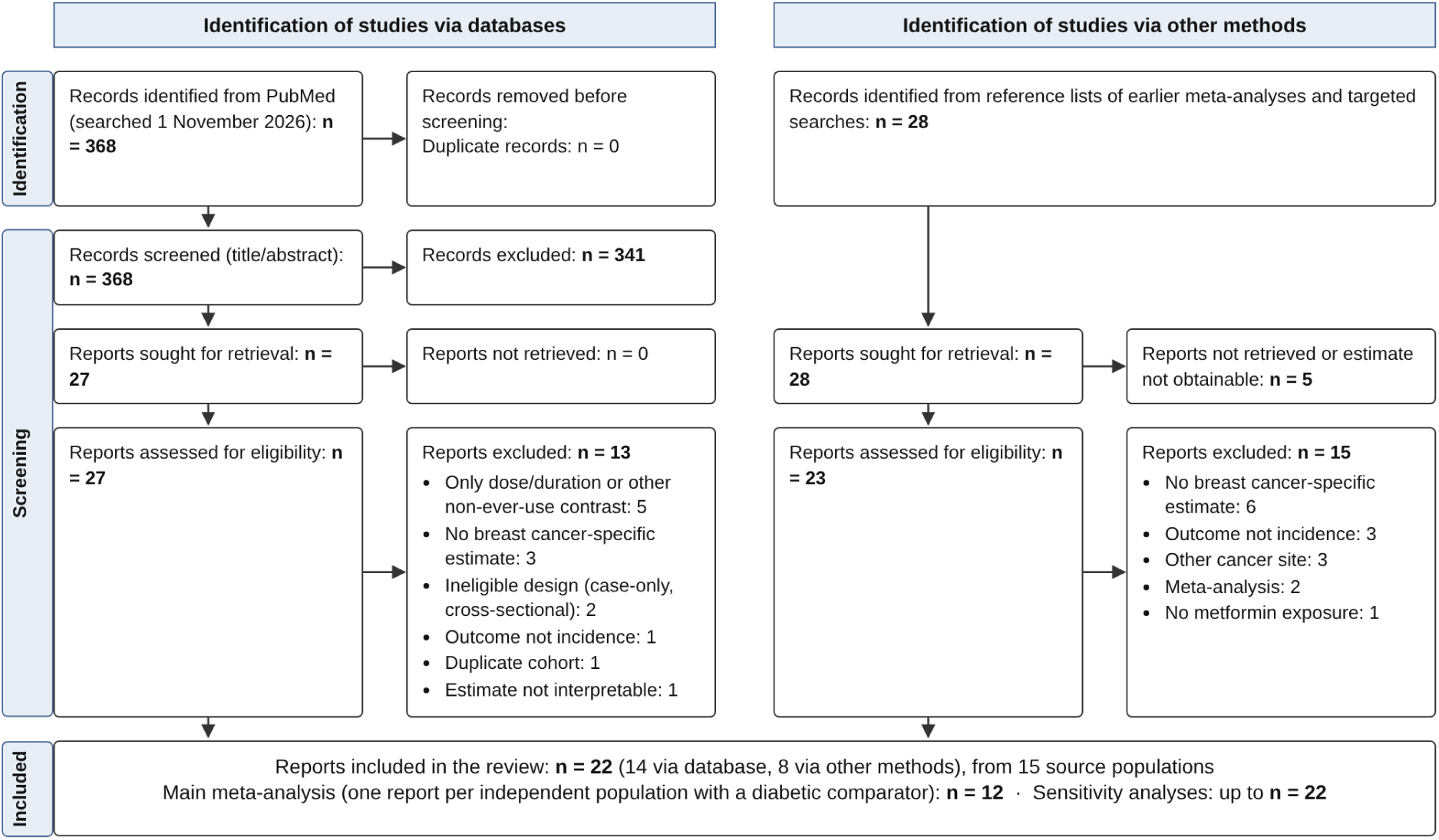
PRISMA 2020 flow diagram. Records and reports identified, screened, excluded and included. The PubMed search was run on 1 November 2026; other methods were reference lists of earlier meta-analyses and targeted searches.

In total, 22 reports from 15 source populations were included (Table 1). They were published between 2009 and 2024 and came from the UK, Denmark, Finland, Germany, the USA, Israel, Taiwan and China. Twelve reports, one per independent source population with a diabetic comparator, formed the main analysis. Nine of the twelve were classified as having addressed time-related biases. Of the other three, two had been identified in earlier methodological work as susceptible to time-related bias [9]: Libby 2009 (time-lag bias from selecting matched non-users with replacement) and Bosco 2011. The third, Tseng 2014, could not be classified from the available text. Four of the twelve used a sulfonylurea comparator, and eight used non-use of metformin among people with diabetes. Three reports with a comparator group that included women without diabetes (Chlebowski 2012, Park 2021 and Rennert 2020) were used only in sensitivity analyses.

**Table 1.** Characteristics of the 22 eligible reports. Estimates are for metformin versus the comparator. Estimates reported as sulfonylurea versus metformin (Hsieh 2012, Qiu 2013, Chen 2015, Zhao 2022) were inverted. ‘Unclear’ was grouped with ‘No’ in the primary subgroup analysis.

| Study | Country / data source | Design | Comparator | Estimate (95% CI) | Time-related bias addressed | Main analysis | PMID / DOI |
|---|---|---|---|---|---|---|---|
| Libby 2009 | UK (Scotland), DARTS/MEMO Tayside | Cohort of new users vs matched never-users | Matched diabetic never-users | HR 0.60 (0.32–1.10) | No | Yes | 19564453 |
| Bosco 2011 | Denmark, Danish registries | Nested case-control | Non-users of metformin | OR 0.81 (0.63–0.96) | No | Yes | 21119073 |
| Tseng 2014 | Taiwan, NHIRD (whole) | Cohort, ever vs never users | Never-users | HR 0.63 (0.60–0.67) | Unclear | Yes | 24816805 |
| Kowall 2015 | Germany/UK, Disease Analyzer | Initiator cohort (ITT); first 12 months excluded | Sulfonylurea initiators | HR 1.03 (0.81–1.30) | Yes | Yes | 26132313 |
| Calip 2016 | USA, Group Health | Cohort with time-varying exposure | Never-users | HR 0.86 (0.65–1.12) | Yes | Yes | 27053250 |
| Hong 2017 | USA, Medicare | Active-comparator new-user cohort | Sulfonylurea initiators | HR 1.2 (0.94–1.6) | Yes | Yes | 28166101 |
| Hosio 2019 | Finland, FinDM | Time-dependent cohort + nested case-control | Non-users | HR 1.02 (0.93–1.11) | Yes | Yes | 30895533 |
| Dankner 2019 | Israel, Clalit | Time-dependent weighted cumulative exposure | No metformin (per 1 DDD/day) | HR 0.88 (0.56–1.39) | Yes | Yes | doi 10.1093/aje/kwz157 |
| Zhao 2022 | China, Yinzhou | New-user active-comparator cohort (MSM-IPW) | Sulfonylurea | HR 1.12 (0.58–2.17) | Yes | Yes | doi 10.3389/fendo.2022.874344 |
| Dickerman 2023 | UK, CPRD | Target trial emulation (ITT) | Non-initiation | HR 1.00 (0.74–1.36) | Yes | Yes | 37227368 |
| Wang 2024 | USA, NHS/NHSII | Cohort with updated exposure and lags | Never-users with T2D | HR 0.92 (0.74–1.15) | Yes | Yes | PMC11096056 |
| Abdallah 2024 | USA, VA | New-user cohort with IPTW | Sulfonylurea new users | HR 0.63 (0.25–1.62) | Yes | Yes | PMC11706726 |
| Bodmer 2010 | UK, GPRD | Nested case-control | No metformin use | OR 1.03 (0.76–1.39) | No | No (sensitivity) | 20299480 |
| Chlebowski 2012 | USA, WHI | Cohort, time-dependent exposure (medication updated at years 1, 3, 6, 9) | Women without diabetes | HR 0.75 (0.57–0.99) | Yes | No (sensitivity) | 22689798 |
| Hsieh 2012 | Taiwan, NHIRD LHID | Cohort analysed by logistic regression | Sulfonylurea monotherapy | OR 0.57 (0.33–0.97) | No | No (sensitivity) | 22719752 |
| Redaniel 2012 | UK, GPRD | Cohort with time-dependent exposure | Sulfonylurea | HR 1.04 (0.79–1.37) | Yes | No (sensitivity) | 22971998 |
| Qiu 2013 | UK, CPRD | Initial monotherapy cohort | Sulfonylurea initiators | HR 0.96 (0.76–1.21) | Yes | No (sensitivity) | 23137378 |
| Tsilidis 2014 | UK, CPRD | Initiator cohort analysed as ITT | Sulfonylurea initiators | HR 1.03 (0.82–1.31) | Yes | No (sensitivity) | 24898303 |
| Chen 2015 | Taiwan, NHIRD LHID | New-onset monotherapy cohort | Sulfonylurea monotherapy | HR 0.80 (0.30–2.13) | Yes | No (sensitivity) | 26357479 |
| Murff 2018 | USA, VA | PS-matched new-user cohort (persistent exposure) | Sulfonylurea new users | HR 1.20 (0.47–2.66) | Yes | No (sensitivity) | 30022336 |
| Park 2021 | USA, Sister Study | Cohort with time-varying T2D and medication | Women without diabetes | HR 0.98 (0.83–1.15) | Yes | No (sensitivity) | PMC7995619 |
| Rennert 2020 | Israel, population-based study (northern Israel) | Population-based case-control | Non-users (with or without diabetes) | OR 0.82 (0.73–0.93) | No | No (sensitivity) | 32446797 |

### Risk of bias

Of the 22 reports, 12 were rated high quality, 8 moderate and 2 low (Table 3). Five reports were rated from the abstract only, because the full text was unavailable to us (Qiu 2013, Tseng 2014, Tsilidis 2014, Kowall 2015 and Rennert 2020). These scored lower partly because the abstract did not report the relevant information. The criteria most often unmet were complete or quantified follow-up (unmet in 14 of 22), adjustment for body mass index (unmet in 10), follow-up of at least 3 years (unmet in 9) and outcome ascertainment by registry or validated codes (unmet in 8). Several active-comparator new-user studies had a median follow-up below 3 years. Two studies previously identified as susceptible to time-related bias, Libby 2009 and Bosco 2011, were rated high quality (8 and 9 of 9).

Restricting the main analysis to the eight high-quality studies gave a pooled RR of 0.96 (0.86–1.07; p = 0.44; I^2^ = 29%). Among these, the six that addressed time-related biases gave 1.02 (0.94–1.10; I^2^ = 0%), and the two that did not (Libby 2009 and Bosco 2011) gave 0.79 (0.64–0.96; p = 0.017).

### Main analysis

The pooled RR for metformin use and incident breast cancer was 0.89 (95% CI 0.74–1.07; p = 0.22; k = 12). Heterogeneity was high: Q = 116.1 (df = 11, p < 0.001), I^2^ = 90.5% and τ^2^ = 0.079. The 95% prediction interval was 0.46–1.72. With REML and the Hartung–Knapp–Sidik–Jonkman correction, the pooled RR was 0.89 (95% CI 0.76–1.03; p = 0.11).

### Subgroup analysis by handling of time-related bias

In the nine studies that addressed time-related biases, the pooled RR was 1.00 (95% CI 0.94–1.08), with no detectable heterogeneity (Q = 5.13, df = 8, p = 0.74; I^2^ = 0%). In the three studies that did not, or were unclear, the pooled RR was 0.69 (95% CI 0.56–0.84; I^2^ = 61%). The difference between the two subgroups was significant (Q = 12.78, df = 1, p = 0.0004; Figure 2). The same pattern appeared when all 19 reports with diabetic comparators were included regardless of overlapping populations: RR 1.00 (0.95–1.07; k = 14; I^2^ = 0%) for studies that addressed time-related biases versus 0.73 (0.59–0.91; k = 5; I^2^ = 73%) for those that did not (between-group Q = 7.31, p = 0.007).

**Figure 2.**
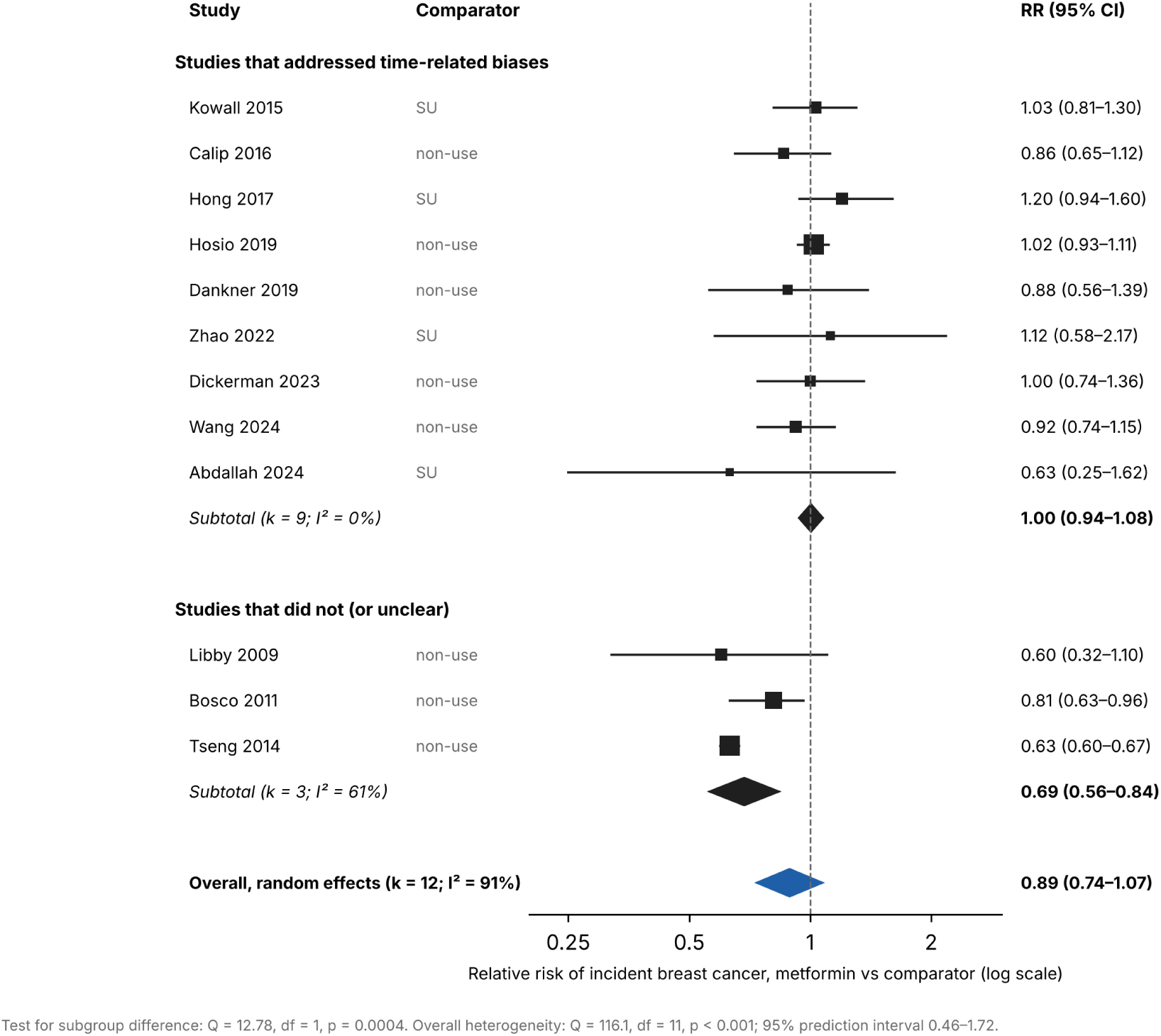
Forest plot of the main analysis by handling of time-related bias. Squares show study estimates, sized by random-effects weight within the subgroup. Diamonds show pooled estimates (DerSimonian–Laird).

This contrast depends on how Tseng 2014 is classified. Its abstract states that metformin was handled as a time-dependent variable [17], but the definition of follow-up start and exposure could not be checked in the full text. If Tseng 2014 is reclassified as having addressed time-related biases, the subgroup that addressed them gives RR 0.92 (0.74–1.14; k = 10; I^2^ = 92%), the other subgroup gives 0.79 (0.64–0.96; k = 2), and the between-group difference is not significant (Q = 1.15, p = 0.28). Under this alternative classification, Tseng 2014 is the only study among the ten that departs from the null; the other nine remain homogeneous (IZ = 0%).

### Other subgroups

With a sulfonylurea comparator, the pooled RR was 1.08 (0.92–1.28; k = 4; I^2^ = 0%). With non-use as the comparator, it was 0.84 (0.67–1.04; k = 8; I^2^ = 93%). The between-group test was Q = 3.38, p = 0.066. All four active-comparator studies had addressed time-related biases, so comparator type and bias handling are not separable in these data. Pooled RRs did not differ by region (Q = 0.76, df = 3, p = 0.86).

### Sensitivity analyses and small-study effects

Omitting Tseng 2014 changed the pooled RR to 0.97 (0.89–1.05) and reduced I^2^ from 90.5% to 10.8%. Omitting any other single study gave pooled RRs between 0.86 and 0.91, with I^2^ of at least 83%. Pooling all 19 reports with diabetic comparators gave 0.91 (0.78–1.05; I^2^ = 87%). Adding the three studies whose comparator included women without diabetes gave 0.90 (0.80–1.02; k = 22). The pooled RR was 0.89 (0.74–1.08) when Tsilidis 2014 replaced Dickerman 2023 for CPRD, 0.91 (0.75–1.09) when Murff 2018 replaced Abdallah 2024 for the VA, and 0.90 (0.74–1.08) when the per-protocol estimate of Dickerman 2023 was used. Egger’s test did not show significant funnel-plot asymmetry (intercept 2.11, SE 1.26; p = 0.13; Figure 3). With 12 studies this test has low power.

**Figure 3.**
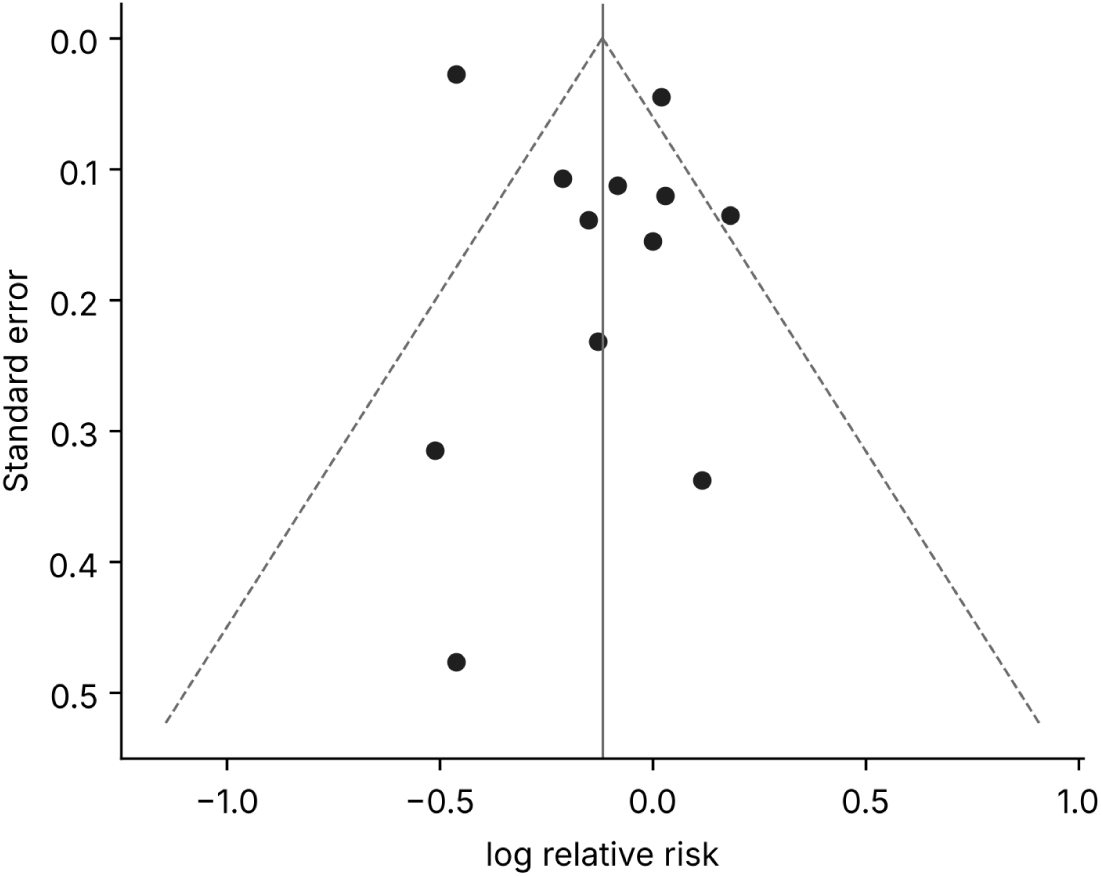
Funnel plot of the 12 studies in the main analysis. Egger’s test: intercept 2.11, SE 1.26, p = 0.13.

### Imeglimin

No eligible clinical study was found, so no meta-analysis of imeglimin could be performed. The PubMed search for “imeglimin” returned 218 records. Combining it with cancer terms returned 10 records, and combining it with “breast” returned 1. None of these was a clinical study reporting cancer incidence. ClinicalTrials.gov listed 10 registered studies of imeglimin: pharmacokinetic, bioequivalence and QT studies, one dose-finding trial, one study of durability of glycaemic control, and one planned comparison with metformin. None had cancer as an outcome. Table 2 lists the evidence that does exist and why it cannot be pooled.

**Table 2.** Available evidence on imeglimin and cancer, and why it cannot contribute to a meta-analysis of breast cancer prevention. For a cohort comparing imeglimin with an active comparator in a 1:1 ratio, detecting an HR of 0.80 with 80% power at a two-sided α of 0.05 would need about 631 breast cancer events [36]. Detecting an HR of 0.90 would need about 2,828 events.

| **Source** | **What it reports** | **Why it cannot be pooled** |
|---|---|---|
| Phase 3 TIMES programme, Japan (>1,100 patients) [12] | HbA1c over 16–52 weeks; adverse events | Treatment duration far shorter than breast cancer latency; breast cancer not reported as an outcome; men included |
| Post-marketing use (approved in Japan, June 2021) | No published cohort reporting cancer outcomes | Around 5 years of use; no comparative study exists |
| Ravichandar & Elango 2024 [24] | T47D and MDA-MB-231 viability (MTT, 48 h) | In vitro; concentration reported without a volume basis; no non-malignant cells; a single study |
| Elango et al. 2024 [25] | Docking of imeglimin derivatives to oestrogen receptor-α | In silico; no effect measure |
| Jiang et al. 2026 [26]; patent US 11,324,752 [27] | Myeloma and hepatocellular carcinoma models at 0.5–25 mmol/L | Not breast tissue; concentrations far above clinical exposure |
| PMDA review, rat 104-week carcinogenicity study [20] | Increased mesenteric lymph node haemangioma in males | Safety signal in animals; not an estimate of human risk |

**Table 3.** Risk of bias (Newcastle–Ottawa Scale) of the 22 included reports. S1–S4, selection; C1, adjustment for age; C2, adjustment for body mass index or obesity; O1–O3 (cohort) or E1–E3 (case-control), outcome or exposure. ★, criterion met; –, not met or not reported. Quality: high 7–9, moderate 4–6, low 0–3. The NOS does not assess time-related bias, which was classified separately (Table 1).

| **Study** | **Design** | **Source** | **S1** | **S2** | **S3** | **S4** | **C1** | **C2** | **O1/E1** | **O2/E2** | **O3/E3** | **Total** | **Quality** |
|---|---|---|---|---|---|---|---|---|---|---|---|---|---|
| Libby 2009 | Cohort | Full text | ★ | ★ | ★ | ★ | ★ | ★ | ★ | – | ★ | 8 | High |
| Bodmer 2010 | Nested case-control | Full text | ★ | ★ | ★ | ★ | ★ | ★ | ★ | ★ | ★ | 9 | High |
| Bosco 2011 | Nested case-control | Full text | ★ | ★ | ★ | ★ | ★ | ★ | ★ | ★ | ★ | 9 | High |
| Chlebowski 2012 | Cohort | Full text | – | ★ | – | ★ | ★ | ★ | ★ | ★ | – | 6 | Moderate |
| Hsieh 2012 | Cohort | Full text | ★ | ★ | ★ | ★ | ★ | – | – | – | – | 5 | Moderate |
| Redaniel 2012 | Cohort | Full text | ★ | ★ | ★ | ★ | ★ | ★ | ★ | – | – | 7 | High |
| Qiu 2013 | Cohort | Abstract | ★ | ★ | – | – | ★ | – | – | – | – | 3 | Low |
| Tseng 2014 | Cohort | Abstract | ★ | ★ | ★ | – | – | – | – | ★ | – | 4 | Moderate |
| Tsilidis 2014 | Cohort | Abstract | ★ | ★ | – | – | – | – | – | – | – | 2 | Low |
| Chen 2015 | Cohort | Full text | ★ | ★ | ★ | ★ | ★ | ★ | – | – | – | 6 | Moderate |
| Kowall 2015 | Cohort | Abstract | ★ | ★ | ★ | – | ★ | – | – | ★ | – | 5 | Moderate |
| Calip 2016 | Cohort | Full text | ★ | ★ | ★ | ★ | ★ | – | ★ | ★ | ★ | 8 | High |
| Hong 2017 | Cohort | Full text | ★ | ★ | ★ | ★ | ★ | ★ | ★ | – | – | 7 | High |
| Murff 2018 | Cohort | Full text | ★ | ★ | ★ | ★ | ★ | ★ | – | – | ★ | 7 | High |
| Hosio 2019 | Cohort | Full text | ★ | ★ | ★ | ★ | ★ | – | ★ | ★ | ★ | 8 | High |
| Dankner 2019 | Cohort | Full text | ★ | ★ | ★ | ★ | ★ | – | ★ | ★ | ★ | 8 | High |
| Park 2021 | Cohort | Full text | – | ★ | – | ★ | ★ | ★ | ★ | ★ | ★ | 7 | High |
| Zhao 2022 | Cohort | Full text | ★ | ★ | ★ | ★ | ★ | ★ | ★ | – | – | 7 | High |
| Dickerman 2023 | Cohort | Full text | ★ | ★ | ★ | ★ | ★ | ★ | ★ | ★ | – | 8 | High |
| Wang 2024 | Cohort | Full text | – | ★ | – | ★ | ★ | ★ | ★ | ★ | – | 6 | Moderate |
| Abdallah 2024 | Cohort | Full text | ★ | ★ | ★ | ★ | ★ | – | – | ★ | – | 6 | Moderate |
| Rennert 2020 | Case-control | Abstract | – | ★ | ★ | – | ★ | – | ★ | ★ | – | 5 | Moderate |

## Discussion

### Principal findings

Across 12 independent populations, metformin use was not associated with a statistically significant reduction in incident breast cancer (pooled RR 0.89, 95% CI 0.74–1.07). The pooled estimate was heterogeneous, and that heterogeneity followed how studies handled time. Nine studies that defined exposure in new users, as time-varying exposure or by target trial emulation gave a pooled RR of 1.00 (0.94–1.08) with I^2^ = 0%. Three studies that did not, or could not be shown to, gave 0.69 (0.56–0.84). A single large ever-versus-never cohort (Tseng 2014) accounted for most of the overall heterogeneity, and the significance of the subgroup difference depends on how that study is classified: without it, I^2^ fell to 10.8% and the pooled RR was 0.97 (0.89–1.05).

### Comparison with earlier work

These results agree with the argument of Suissa and Azoulay that the large reductions reported in early observational studies are explained by immortal time and time-window bias [9]. They also agree with the null result of the target trial emulation in CPRD [16] and with the null GRADE-assessed meta-analysis of Tang et al. [6]. Meta-analyses that reported protection pooled studies without separating them by design [5, 7]. The MA.32 trial addressed recurrence in non-diabetic women and does not test primary prevention [10]. Taken together, the observational evidence does not support a clinically relevant preventive effect of metformin on breast cancer in women with diabetes. It also does not exclude a modest effect in specific subgroups, such as long-term users or women with marked hyperinsulinaemia, because few studies reported these strata in a comparable way.

### How time-related biases produce an apparent protective effect

The biases at issue share one feature: person-time or exposure opportunity is allocated unequally between the groups being compared, and the error almost always favours the drug.

- **Immortal time bias.** In a cohort study, time from cohort entry to the first metformin prescription is event-free by definition, because a patient must survive without cancer to become a user. When this time is counted as exposed, the person-time denominator of users is inflated and that of non-users is reduced [8, 31]. In a worked example by Suissa and Azoulay, rates were 10.2 per 10,000 person-years in users and 23.2 in non-users (rate ratio about 0.44). After 16,616 immortal person-years were reassigned to the unexposed group, the rates became 12.4 and 10.7 (rate ratio about 1.16) [9]. An apparent 56% reduction became a 16% increase. Classifying women as ever-users or never-users over the whole follow-up, as in Tseng 2014 [17], is susceptible to this bias unless exposure is modelled as time-varying from a common start.
- **Time-window bias.** In case-control studies, exposure is often measured over different lengths of time for cases and controls. The longer the window, the more prescriptions accumulate, so cumulative or long-term categories of use can be misclassified. This bias has been described for the nested case-control studies of Bodmer 2010 and Bosco 2011 [9].
- **Time-lag bias.** The compared groups may sit at different stages of the disease, or they may be selected in a way that shifts events across time zero. In Libby 2009, matched non-users were selected with replacement. As a result, cancers that should have excluded a non-user were counted as outcomes after the index date, which inflated the cancer rate in the non-user group [9].
- **Detection bias and reverse causation.** Initiating a drug changes contact with health care, so cancers may be found earlier or later than they would otherwise be. In a cohort of new users, the HR for breast cancer was 0.66 (0.43–1.02) in the first 3 months after metformin initiation and 1.00 (0.98–1.02) thereafter [32].

These biases can be avoided by design:

- aligning eligibility, treatment assignment and the start of follow-up at a common time zero, as in target trial emulation [33];
- comparing new users of metformin with new users of an alternative glucose-lowering drug (active-comparator, new-user design) [34, 35];
- modelling exposure as time-varying, so that pre-treatment time counts as unexposed [31];
- applying a lag, or excluding the first 6–12 months of follow-up;
- in case-control studies, measuring exposure over windows of equal length for cases and controls.

In this review, studies that used one or more of these approaches gave a pooled RR of 1.00 (0.94–1.08), whereas studies that did not gave 0.69 (0.56–0.84). The size of the protective associations reported earlier is therefore within the range that these biases alone can produce. Because the significance of the subgroup difference depends on how Tseng 2014 is classified, this comparison should be read together with the sensitivity analysis. For future pharmacoepidemiological work, including any study of imeglimin, these design features should be specified in advance rather than checked after the analysis.

Generic quality scales do not detect these biases. Libby 2009 and Bosco 2011 scored 8 and 9 of 9 on the NOS, but both had been shown to be susceptible to time-related bias [9]. When the analysis was restricted to high-quality studies, the contrast between studies that addressed time-related bias (RR 1.02) and those that did not (RR 0.79) persisted. Restricting by NOS score alone would therefore not have removed the source of the apparent protection, and design features that determine time zero and exposure classification need to be assessed explicitly.

### Imeglimin: no clinical evidence, and what the structure does and does not imply

We found no clinical study, observational or randomised, that reports breast cancer incidence in imeglimin users. At present, the effect of imeglimin on breast cancer risk therefore cannot be evaluated. The following structure-based considerations are hypotheses for future work, not evidence of benefit.

**Why a meta-analysis of imeglimin is not yet possible.** Four obstacles stand in the way, and none can be overcome by statistical methods.

1. **There is nothing to pool.** No clinical study has reported breast cancer incidence in imeglimin users, so the number of eligible studies is zero.
2. **Exposure time is too short.** Imeglimin was approved in Japan in June 2021. The phase 3 trials lasted 16–52 weeks [12], and post-marketing use spans about 5 years. Breast cancer develops over many years, and a preventive effect, if any, would be expected only after long exposure.
3. **The non-clinical data cannot be combined with clinical data or with each other.** A cell viability IC50, a docking score and a hazard ratio measure different quantities, and there is only one in vitro study in breast cancer cells [24]. Placing these on a common scale would produce a number with no interpretation.
4. **The events required are large.** A comparative cohort would need about 631 breast cancer events to detect an HR of 0.80, and about 2,828 to detect an HR of 0.90. The observational studies of metformin that addressed time-related biases are consistent with no effect (RR 1.00, 0.94–1.08), and imeglimin acts more weakly than metformin on the pathways proposed for metformin (below). An effect stronger than an HR of 0.80 is therefore implausible, and the required number of events is likely to be at the upper end of this range.

The realistic next step is therefore a single, adequately powered, new-user active-comparator cohort in Japanese claims data, not a meta-analysis, with follow-up long enough for breast cancer latency.

#### Shared scaffold, different conformation

Imeglimin is (6R)-N2,N2,6-trimethyl-3,6-dihydro-1,3,5-triazine-2,4-diamine, given as the hydrochloride (MW 191.66) [20]. Like metformin, it carries an N,N-dimethylamino group on a guanidine-derived, nitrogen-rich core. In imeglimin the biguanide-like chain is closed into a dihydrotriazine ring by a carbon that bears a methyl group, which also creates a stereocentre. Both molecules are small, polar organic cations. Consistent with this, imeglimin is a substrate of OCT1, OCT2, MATE1 and MATE2-K and is excreted largely unchanged in urine [21], a disposition similar to that of metformin. One structural expectation is therefore that imeglimin, like metformin, would enter breast epithelial or tumour cells only to the extent that these cells express organic cation transporters. Breast cancer cell lines differ widely in OCT1 and OCT3 expression [28], and in xenografts the effect of metformin tracked intratumoural drug concentration and cation-transporter expression [29]. Whether imeglimin reaches breast tissue at all has not been measured.

#### Different action at the respiratory chain

The ring closure is associated with a different mitochondrial profile. Imeglimin partially inhibits complex I, restores deficient complex III activity, lowers reactive oxygen species and prevents opening of the mitochondrial permeability transition pore [11]. In endothelial cells it did this without lowering oxygen consumption [22]. Imeglimin activates AMPK in hepatocytes, but less potently than metformin and only at 0.25–10 mmol/L [23]. The direct anticancer mechanism proposed for metformin depends on energetic stress through complex I inhibition, AMPK activation and mTOR suppression [3]. A drug designed to cause less energetic stress would be expected to act less strongly through this route, not more.

#### Systemic effects point in different directions

The indirect mechanism proposed for metformin is lower circulating insulin [3]. Imeglimin increases glucose-stimulated insulin secretion [11], so the insulin-lowering hypothesis does not transfer to it. Lower oxidative stress could in principle reduce oxidative DNA damage in mammary epithelium, but this has not been tested. Imeglimin also raises NAD⁺ through the salvage pathway [11]; NAMPT, the rate-limiting enzyme of that pathway, is overexpressed in breast cancer and is itself an anticancer drug target [30], so the direction of this effect on breast carcinogenesis is uncertain.

#### Available experimental data

Imeglimin reduced viability of T47D and MDA-MB-231 cells in an MTT assay. That study reported concentrations without a volume basis and tested no non-malignant cells [24]. A docking study of imeglimin derivatives against oestrogen receptor-α was entirely in silico [25]. In multiple myeloma cells, imeglimin inhibited proliferation and oxidative phosphorylation at 0.5–5 mmol/L [26], and a patent reports hepatocellular carcinoma cell inhibition at 2.5–25 mmol/L [27]. For comparison, steady-state Cmax at 500–2000 mg twice daily in Japanese subjects was 994–2267 ng/mL as the hydrochloride [21], about 5–12 µmol/L. The lowest concentration with an anticancer effect is therefore at least about 40-fold above clinical exposure.

#### Safety signal

In the 104-week rat carcinogenicity study, mesenteric lymph node haemangioma increased in males: 10 (negative control), 3 (vehicle control), and 8, 11 and 18 at 125, 250 and 500 mg/kg/day. The 26-week Tg-rasH2 mouse study and genotoxicity studies were negative [20]. Any chemoprevention hypothesis has to be weighed against this finding.

#### What would be needed

Experiments at clinically attainable concentrations in mammary epithelial and breast cancer cells with characterised OCT expression; measurement of drug concentrations in breast tissue; and, because imeglimin was first approved in Japan, a new-user active-comparator cohort in Japanese claims data (for example versus DPP-4 inhibitors) with sufficient follow-up for breast cancer latency.

### Strengths and limitations

The report deviates from the registered protocol in scope and search sources (see Methods), and the subgroup analysis by handling of time-related bias was defined during the re-analysis rather than in the protocol. We have therefore reported every analysis we performed, including those whose results depend on study classification. Only one bibliographic database (PubMed) was searched, and titles and abstracts were screened by one reviewer. To reduce the risk of missing studies, we also screened the reference lists of earlier meta-analyses and searched specifically for cancer-incidence studies that report breast cancer as one of several sites; eight of the 22 included reports were found only this way. Strengths are the restriction to primary studies with breast-specific estimates, the handling of overlapping databases, and the comparison by time-related bias, which explained the heterogeneity. Limitations follow. The second extraction was performed by a large language model rather than a second human reviewer, although every value used was verified against the source by a human author. The full texts of four reports (Qiu 2013, Tseng 2014, Tsilidis 2014, Kowall 2015) were not available to us, so their estimates were taken from the abstracts. Five further reports could not be assessed. Risk of bias was scored by one reviewer with human verification, and five reports were scored from the abstract alone, which tends to understate their quality. The time-bias classification of one large study (Tseng 2014) could not be confirmed from the available text, and the significance of the subgroup difference depends on it. Comparator type and bias handling were confounded, because every active-comparator study had addressed time-related bias. Follow-up in several new-user studies was short relative to breast cancer latency. HRs and ORs were pooled as approximations of a common RR. Data on menopausal status, tumour subtype, dose and duration were too sparse to pool.

## Conclusions

In studies designed to avoid time-related biases, metformin use was not associated with incident breast cancer (RR 1.00, 95% CI 0.94–1.08). The protective associations reported earlier come from studies susceptible to these biases and from one large Taiwanese cohort whose handling of exposure time could not be verified. For imeglimin, no clinical data on breast cancer exist, so a meta-analysis cannot be performed: there are no eligible studies, exposure since approval spans only about 5 years, and the available non-clinical results cannot be combined with clinical estimates. Its structural kinship with metformin does not carry over to the mechanisms proposed for metformin’s anticancer action, and any preventive potential remains a hypothesis that requires both experimental and pharmacoepidemiological testing.

## Declarations

### Ethics approval

Not required. This study analysed published aggregate data only.

### Data and code availability

Supplementary Data contain the extraction table (with the source location of every estimate), the extraction form and instructions used for the second extraction, the record of discrepancies and their resolution, the PubMed search record and list of retrieved PMIDs, the list of excluded reports, the risk-of-bias assessment with supporting text, and the Python analysis code.

### Use of generative AI

Large language models were used as follows. Claude (Anthropic) assisted with literature retrieval, data extraction, statistical analysis code and drafting. ChatGPT (OpenAI) performed an independent second data extraction. All values used in the analyses were verified against the original reports by a human author (KO), and the authors take full responsibility for the content.

### Author contributions

Conceptualization: K.O. Methodology: K.O. Investigation: M.A. and K.O. Data curation and verification: K.O. Formal analysis: K.O. Visualization: K.O. Writing — original draft: K.O. Supervision and project administration: K.O. Y.H., H.H., M.M. and Y.M. advised on study design and reviewed the manuscript. All authors approved the final version.

### Preprint server note

This work concerns human health outcomes and is intended for medRxiv rather than bioRxiv.

## Data Availability

All data used in this study were extracted from previously published articles, which are cited in the manuscript and listed with their PMIDs/DOIs in Table 1. The full extraction table (with the source location of every estimate), the PubMed search record and list of retrieved PMIDs, the risk-of-bias assessment, the list of excluded reports, and the Python analysis code are provided as Supplementary Data.

The primary studies in Table 1 are cited there by PMID or DOI.

